# Clinical Trends Among U.S. Adults Hospitalized with COVID-19, March-December 2020

**DOI:** 10.1101/2021.04.21.21255473

**Authors:** Shikha Garg, Kadam Patel, Huong Pham, Pam D. Kirley, Breanna Kawasaki, Kimberly Yousey-Hindes, Evan J. Anderson, Andrew Weigel, Patricia A. Ryan, Libby Reeg, Kathryn Como-Sabetti, Sarah Shrum Davis, Alison Muse, Nancy M. Bennett, Laurie Billing, Melissa Sutton, H. Keipp Talbot, Mary Hill, Jonathan Wortham, Lindsay Kim, Fiona Havers, COVID-NET Surveillance Team

## Abstract

**Background:** The COVID-19 pandemic has caused substantial morbidity and mortality.

**Objectives:** To describe monthly demographic and clinical trends among adults hospitalized with COVID-19.

**Design:** Pooled cross-sectional.

**Setting:** 99 counties within 14 states participating in the Coronavirus Disease 2019-Associated Hospitalization Surveillance Network (COVID-NET).

**Patients:** U.S. adults (aged ≥18 years) hospitalized with laboratory-confirmed COVID-19 during March 1-December 31, 2020.

**Measurements:** Monthly trends in weighted percentages of interventions and outcomes including length of stay (LOS), intensive care unit admissions (ICU), invasive mechanical ventilation (IMV), vasopressor use and in-hospital death (death). Monthly hospitalization, ICU and death rates per 100,000 population.

**Results:** Among 116,743 hospitalized adults, median age was 62 years. Among 18,508 sampled adults, median LOS decreased from 6.4 (March) to 4.6 days (December). Remdesivir and systemic corticosteroid use increased from 1.7% and 18.9% (March) to 53.8% and 74.2% (December), respectively. Frequency of ICU decreased from 37.8% (March) to 20.5% (December). IMV (27.8% to 8.7%), vasopressors (22.7% to 8.8%) and deaths (13.9% to 8.7%) decreased from March to October; however, percentages of these interventions and outcomes remained stable or increased in November and December. Percentage of deaths significantly decreased over time for non-Hispanic White patients (p-value <0.01) but not non-Hispanic Black or Hispanic patients. Rates of hospitalization (105.3 per 100,000), ICU (20.2) and death (11.7) were highest during December.

**Limitations:** COVID-NET covers approximately 10% of the U.S. population; findings may not be generalizable to the entire country.

**Conclusions:** After initial improvement during April-October 2020, trends in interventions and outcomes worsened during November-December, corresponding with the 3^rd^ peak of the U.S. pandemic. These data provide a longitudinal assessment of trends in COVID-19-associated outcomes prior to widespread COVID-19 vaccine implementation.

## Introduction

The Centers for Disease Control and Prevention (CDC) estimates that 83.1 million total infections, 70.4 million symptomatic illnesses and 4.1 million hospitalizations associated with coronavirus disease 2019 (COVID-19) occurred in the United States as of December 2020 (1, 2). The clinical epidemiology of COVID-19 among U.S. adults has been well described, with older age and underlying conditions identified as risk factors for COVID-19-associated hospitalization and mortality (3-10). However, data on trends in clinical characteristics and outcomes of COVID-19-associated hospitalizations (11-16) are limited, and few studies to date include data from the 3^rd^ peak of the U.S. pandemic. CDC’s Coronavirus Disease 2019-Associated Hospitalization Surveillance Network (COVID-NET) (17) has shown that 25% of COVID-19-associated hospitalizations during March-December 2020 required intensive care unit (ICU) admission, 14% received invasive mechanical ventilation, and 11% died in-hospital (<u>COVID-19 Hospitalizations (cdc.gov)</u>). However, cumulative data do not describe changing trends over time, which may be impacted by changes in COVID-19 epidemiology (18), implementation of mitigation measures (19, 20), evolving COVID-19 treatments (21-24), and knowledge, experience and capacity of healthcare providers and systems caring for hospitalized patients with COVID-19 (12, 14-16). Using COVID-NET data, we described monthly trends in clinical characteristics, interventions and outcomes among adults hospitalized with COVID-19 during March–December 2020, prior to the large-scale availability of COVID-19 vaccines.

## Methods

COVID-NET conducts population-based surveillance for laboratory-confirmed COVID-19– associated hospitalizations among persons of all ages in 99 counties in 14 states participating in the Emerging Infections Program (California, Colorado, Connecticut, Georgia, Maryland, Minnesota, New Mexico, New York, Oregon, Tennessee) and the Influenza Hospitalization Surveillance Project (Iowa, Michigan, Ohio, Utah). COVID-NET covers a catchment population of approximately 32 million persons (∼10% of the U.S. population) and collects data from over 250 acute care hospitals (17). Hospitalized patients who are residents of the surveillance catchment area and have SARS-CoV-2 detected by positive molecular or rapid antigen testing during hospitalization or within 14 days before admission are included in COVID-NET. SARS-CoV-2 testing is performed at the discretion of health care providers or according to hospital testing policies.

A minimum set of data are collected on all identified cases to produce weekly hospitalization rates (<u>COVID-19 Hospitalizations (cdc.gov)</u>). Incidence rates are calculated using the National Center for Health Statistics’ vintage 2019 bridge-race postcensal population estimates for the counties included in surveillance (25). Detailed clinical data are collected for a random sample of cases aged ≥18 years stratified by age and surveillance site. Random numbers are auto generated and assigned to each case upon entry into the surveillance database. Trained surveillance staff conduct medical chart abstractions using a standardized case report form. Data on race and ethnicity were categorized as follows: non-Hispanic White (White), non-Hispanic Black (Black), Hispanic or Latino (Hispanic), non-Hispanic Asian or Pacific Islander (Asian/PI), non-Hispanic American Indian or Alaska Native (AI/AN) and people of more than one race/ethnicity. If ethnicity was unknown (3.8% of cases), non-Hispanic ethnicity was assumed. Facility residence at admission was defined as residence in rehabilitation facilities, assisted living/residential care, group homes, nursing homes, skilled nursing facilities, long-term care facilities, long term acute care hospitals, alcohol/drug treatment centers, and psychiatric facilities. Healthcare workers were defined as all paid and unpaid persons serving in healthcare settings who have the potential for direct or indirect exposure to patients or infectious materials. Patients were defined as having 0, 1, 2, or ≥3 underlying condition categories rather than individual conditions as follows: chronic lung disease (including asthma), chronic metabolic disease (including diabetes mellitus), cardiovascular disease (excluding hypertension), blood disorders, neurologic disorders, immunosuppressive conditions, chronic renal disease, rheumatologic/autoimmune conditions, hypertension, obesity (defined as body mass index ≥30 kg/m^2^), feeding-tube dependent, ventilator-dependent and wheel-chair-dependent. Any medication prescribed for treatment of SARS-CoV-2, including investigational agents or treatment through a clinical trial, was classified as a COVID-19–associated treatment. Invasive mechanical ventilation, bilevel positive airway pressure (BiPAP), continuous positive airway pressure (CPAP), and high-flow nasal cannula (HFNC) were defined based on the highest level of respiratory support received. Vasopressor use was included only if administered as a continuous infusion. Renal replacement therapy (RRT) during hospitalization was described regardless of whether a patient received RRT prior to hospitalization. Discharge disposition was collected, including whether a patient was discharged from the hospital or died in-hospital.

Data from all cases aged ≥18 years and hospitalized with COVID-19 during March 1–December 31, 2020 were used to describe patient demographics and hospitalization rates. All other analyses were limited to sampled adult patients hospitalized with COVID-19 for whom medical chart abstractions were completed and a discharge disposition was known; weights were applied to reflect the probability of being sampled for medical chart abstraction. Sample sizes were generated to allow for monthly estimation of the prevalence of clinical parameters by 3 adult age groups (18-49 years, 50-64 years and ≥65 years), and separately by 3 racial/ethnic groups (White, Black and Hispanic persons). Due to small sample sizes, we were unable to generate monthly estimates for other racial/ethnic groups. Sample sizes were generated to produce relative standard errors (RSE) <0.3 for clinical parameters with prevalence estimates ≥10% (all estimates with RSE >0.3 are noted in supplemental tables). Descriptive statistics were generated overall and by month from March through December 2020. To produce robust prevalence estimates stratified by both age and race/ethnicity, data from multiple months were combined (i.e., March-May, June-September, October-December). For all analyses using sampled data, weighted percentages and unweighted case counts are presented. Logistic regression models (for all binary measures) were used to test the statistical significance of monthly trends by assigning MMWR month (or groups of months as indicated) as a single continuous predictor in models for all ages combined and for each individual age group strata or age by race strata. The models assumed overall linear effects for outcomes measured. Median length of stay (LOS) (for patients who were discharged alive or died in-hospital) and days from symptom onset to admission were compared across months using the Mann-Whitney-U test. P-values <0.05 were considered statistically significant. We calculated unadjusted hospitalization incidence rates per 100,000 population by taking the total number of cases each month overall and within each age group, divided by the National Center for Health Statistics’ vintage 2019 bridge-race postcensal population estimates for the counties included in surveillance. Incidence rates for ICU admissions and deaths among hospitalized patients were calculated similarly, instead using weighted numbers of sampled cases as the numerators. Taylor series linearization method was used for variance estimation. All analyses were conducted using SAS 9.4 software (SAS Institute Inc., Cary, NC).

This activity was reviewed by CDC and was conducted consistent with applicable federal law and CDC policy (see e.g., 45 C.F.R. part 46.102(l)(2), 21 C.F.R. part 56; 42 U.S.C. §241(d); 5 U.S.C. §552a; 44 U.S.C. §3501 et seq.) Sites participating in COVID-NET obtained approval from their respective state and local Institutional Review Boards, as applicable.

## Results

### Hospitalization Rates and Demographic Characteristics of all Adults

During March 1–December 31, 2020, among adults aged ≥18 years, hospitalization rates ranged from 20.2 per 100,000 population (March) to 105.3 (December); rates were highest among adults aged ≥65 years, ranging from 45.4 (March) to 293.8 (December) (Supplemental Table 1). Among the 116,743 adult cases, 26.4% were 18-49 years, 27.7% were 50-64 years and 46.0% were ≥65 years of age. The median age was 62.0 years (IQR 48.1-74.6); while the median age initially decreased from 60.9 years in March to 55.6 years in June, it increased in subsequent months to 65.4 years in December (Table 1). Overall, 50.7 % of cases were male with minimal variation in sex distribution over time; 40.8% of cases were White, 26.9% were Black, and 20.0% were Hispanic.

**Table 1.**
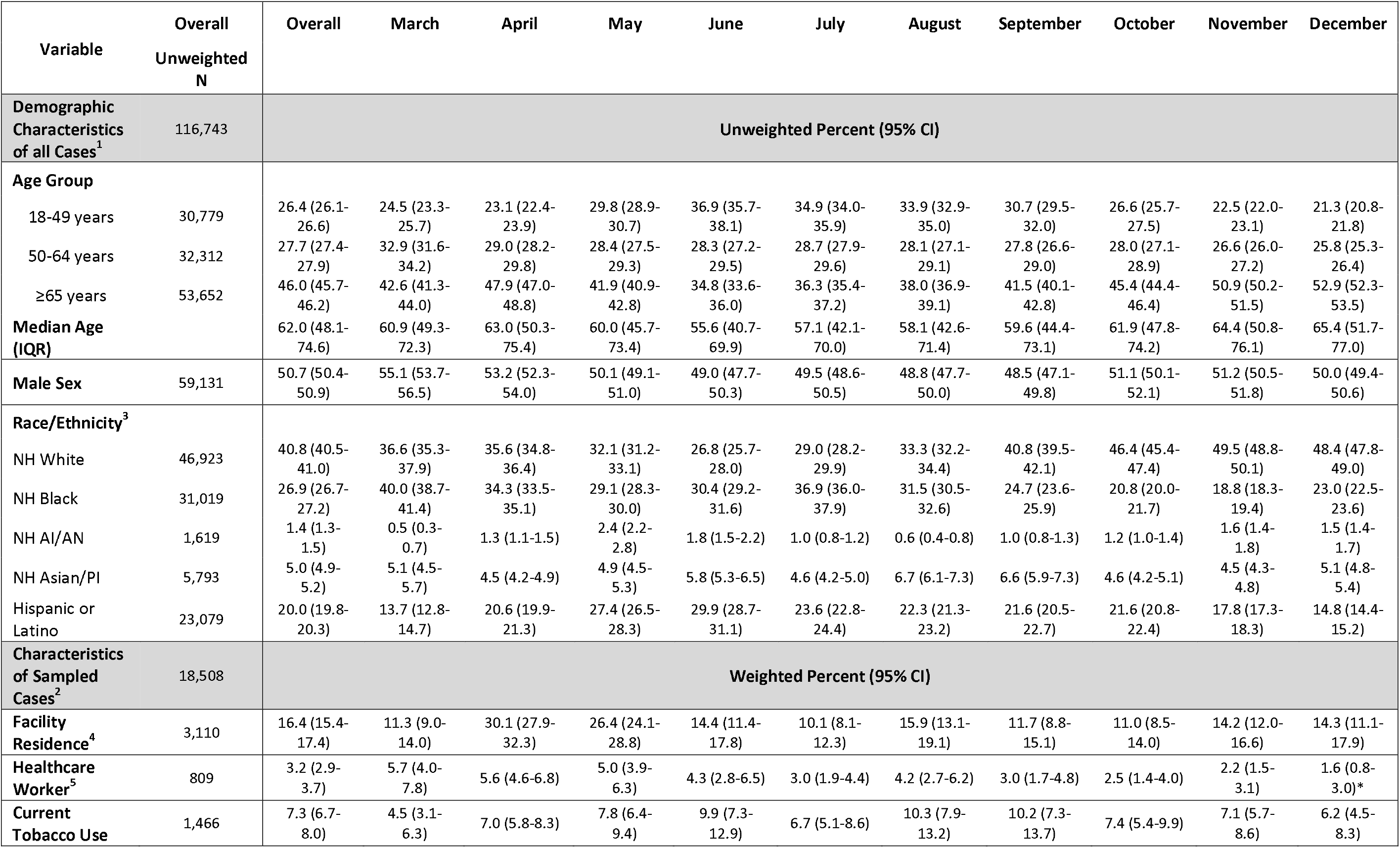
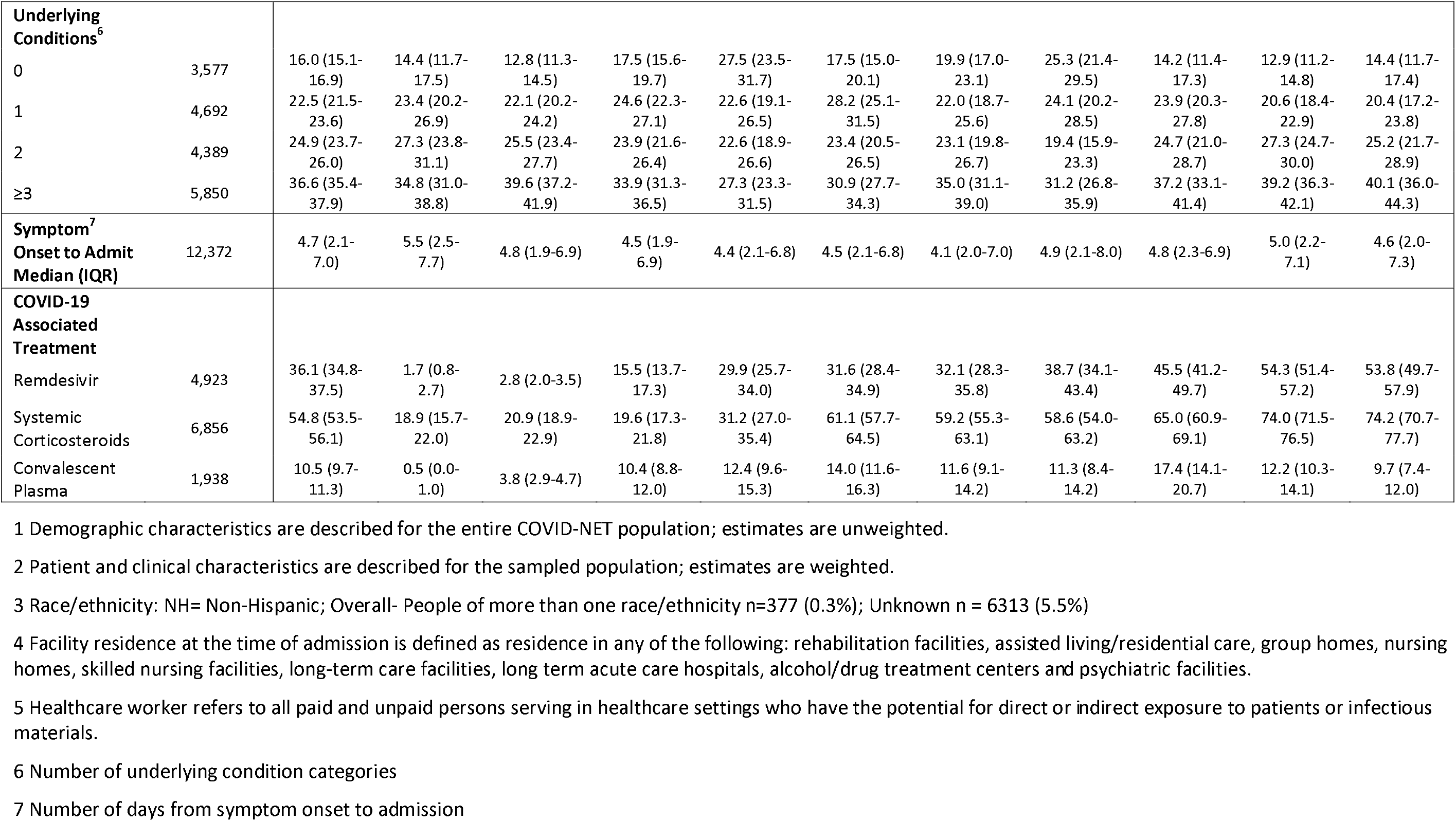
Demographic and Clinical Characteristics of Adults with Laboratory-Confirmed COVID-19-Associated Hospitalizations by Month, COVID-NET, March-December 2020.

### Clinical Characteristics and COVID-19-Associated Treatments among a Sample of Adults

Chart reviews were completed, and discharge dispositions ascertained on a sample of 18,508 adult hospitalized cases. Overall, demographic characteristics of sampled cases were similar to that of all hospitalized cases (Supplemental Table 2). The percentage of cases admitted from a facility varied from 10.1% to 30.1%, without a clear trend over time (Table 1). The percentage of cases identified as healthcare workers decreased from 5.7% in March to 1.6% in December. Most cases had at least 2 underlying condition categories, with small variations over time. However, use of COVID-19-associated treatments varied significantly; remdesivir use increased from 1.7% to 53.8% and systemic corticosteroid use increased from 18.9% to74.2% from March to December (Table 1). While ≥ 40% of patients received hydroxychloroquine treatment in March and April, ≤1% received it after May 2020 (Supplemental Table 3).

### Length of Hospital Stay, Interventions and Outcomes among a Sample of Adults

The median LOS for all patients (including those discharged alive and those who died in-hospital) significantly decreased from 6.4 days (IQR 2.9-16.8) in March to 4.7 days (IQR 2.2-9.2) in July, then remained relatively unchanged from July through December (Figure 1A)). Trends in median LOS varied by age group. The median LOS was 4.6 days (IQR 2.3-8.8) for patients who were discharged alive and 10.4 days (IQR 5.0-18.4) for those who died in-hospital (data not shown).

**Figure 1.**
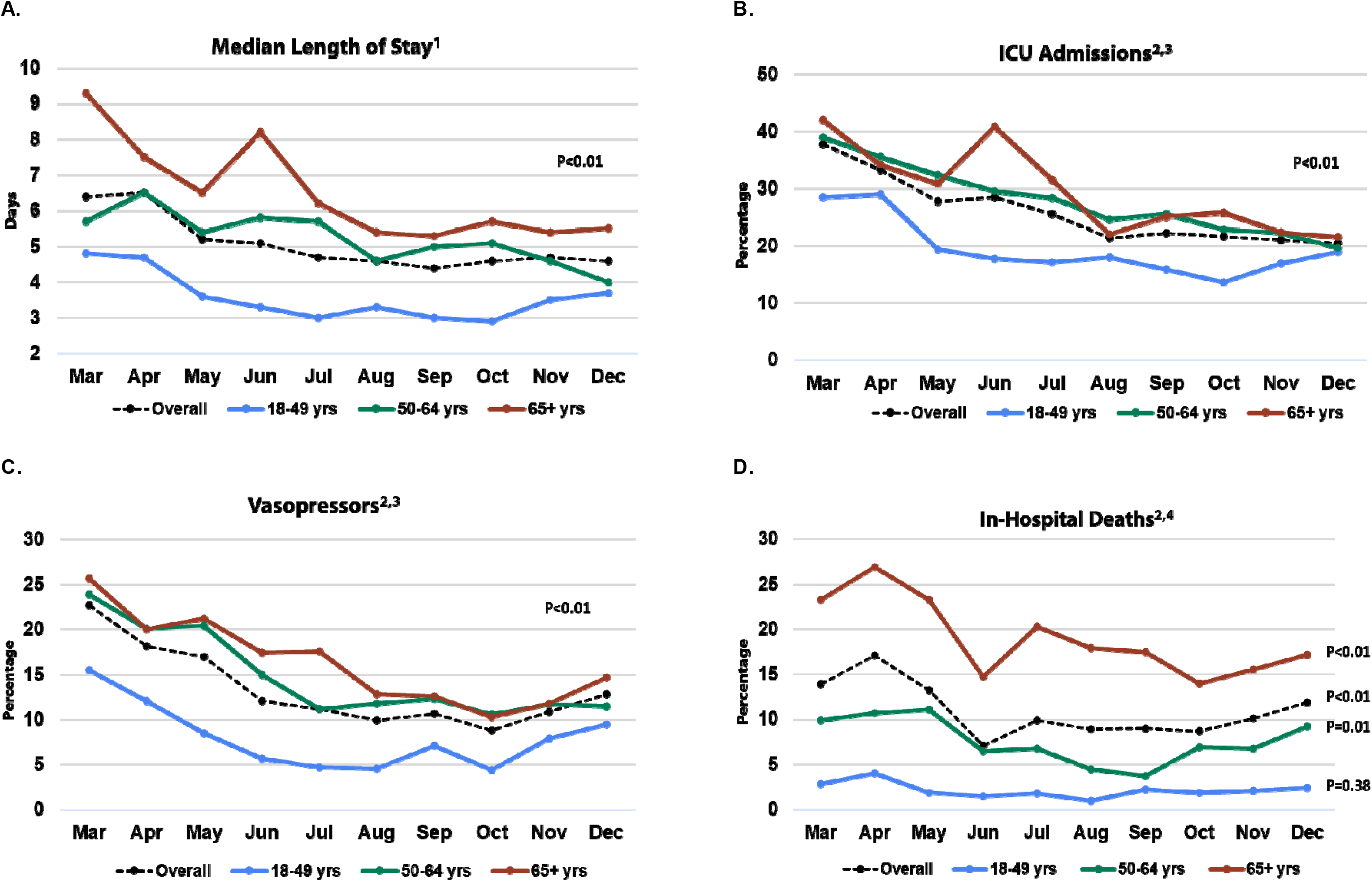
Trends in Length of Stay and Percentages of Interventions and Outcomes among Hospitalized Adults with COVID-19 by Age Group and Month, COVID-NET, March-December 2020. 1. Median lengths of stay for all patients (including those who died in-hospital and were discharged alive) were compared across months using the Mann-Whitney-U test; p-value <0.01 for all ages combined and each individual age group 2. Logistic regression models used to test statistical significance of monthly trends for ICU, vasopressors and death by assigning month as a single continuous predictor in models for all ages combined and individual age group. 3. P-value <0.01 for all ages combined and each individual age group 4. P-value ≤0.01 for all ages combined and for adults 50-64 years and ≥65 years of age

The percentage of hospitalized adults with ICU admission significantly decreased from 37.8% (March) to 20.5% (December) (Figure 1B). ICU admissions significantly decreased for all age groups. Vasopressor use decreased from 22.7% (March) to 12.8% (December) (Figure 1C) with similar trends across the different age groups. Overall, RRT use did not significantly change over time and ranged from 4.1% to 6.7% (Supplemental Table 4).

Overall, 11.4% (95%CI 10.5-12.2) of patients died in-hospital; case fatality was 2.2% (95%CI 1.7-2.8) among adults 18-49 years, 8.0% (95%CI 6.9-9.3) among adults 50-64 years and 18.8% (95%CI 17.1-20.5) among adults ≥65 years of age (data not shown). In-hospital deaths for all ages combined decreased over time, ranging from 13.9% (March) to 11.9% (December) (Figure 1D). Among adults aged ≥65 years, in-hospital deaths decreased from a high of 26.9% (April) to a low of 14% (October) but increased to 17.2% in December. Among adults aged 50-64 years, in-hospital deaths initially decreased from a peak of 10.7% (April) to a low of 3.7% (September) but increased to 9.2% (December). Among all patients who were discharged alive, 3.1% (95%CI 2.5-3.8%) were discharged to hospice (range: 1.2 % in March to 4.1% in December) (data not shown). Among adults aged ≥65 years, 6.7% (95%CI 5.4-8.3%) were discharged to hospice.

### Highest Level of Respiratory Support Received among a Sample of Adults

The percentage of patients receiving invasive mechanical ventilation as the highest level of respiratory support significantly decreased from 27.8% (March) to 12.3% (December) (Figure 2A); trends were similar across all 3 age groups (Figure 2B). The percentage of patients receiving HFNC significantly increased from 5.6% (March) to 8.7% (December) but varied over time; the percentage of patients receiving BIPAP or CPAP significantly increased from 2.5% (March) to 7.5% (December). While monthly changes in HFNC were not significant when stratified by the 3 age groups (Figure 2C), there were significant increases in BIPAP or CPAP use over time across all 3 age groups (Figure 2D).

**Figure 2.**
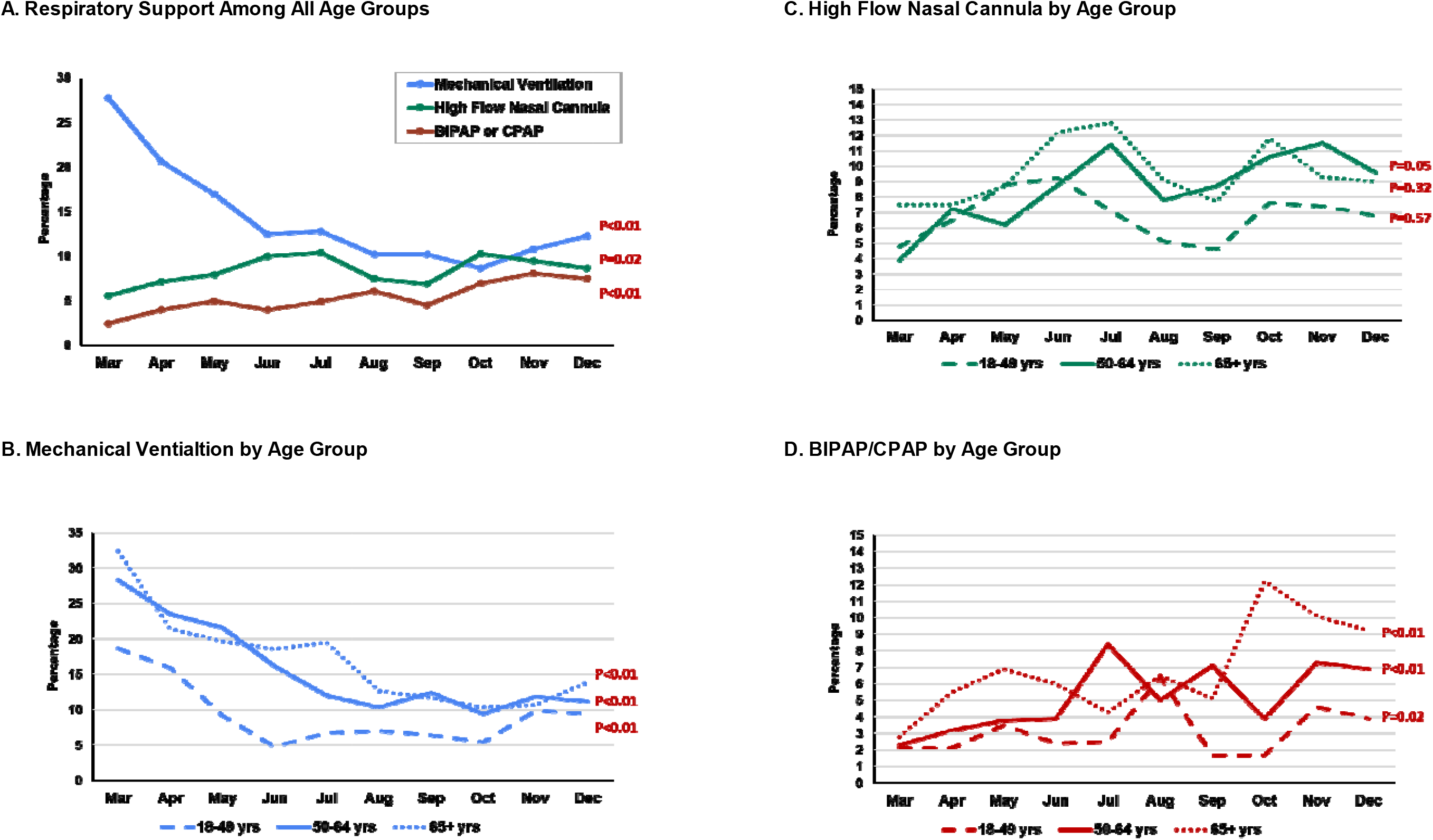
Trends^1^ in Highest Level of Respiratory Support Received among Hospitalized Adults with COVID-19 by Month, COVID-NET, March-December 2020. 1 Logistic regression models were used to test the statistical significance of monthly trends in interventions by assigning MMWR month as a single continuous predictor in models for all ages combined and for each individual age group strata

### Trends in Interventions and Outcomes by Race/Ethnicity and Age Group for three time periods: March-May, June-September, October-December

Among patients 18-49 years and ≥65 years of age, percentage of ICU admissions significantly decreased over time for White and Black patients, but not for Hispanic patients (Figure 3A and 3C, Supplemental Table 5). For patients 50-64 years of age, percentage of ICU admissions significantly decreased over time for all 3 racial/ethnic groups (Figure 3B, Supplemental Table 5). Among White patients, there was a significant decrease in percentage of in-hospital deaths over time for all 3 age groups (Figure 3D-F, Supplemental Table 5); however, there was no significant decrease in percentage of in-hospital deaths among Black or Hispanic patients across any age group.

**Figure 3.**
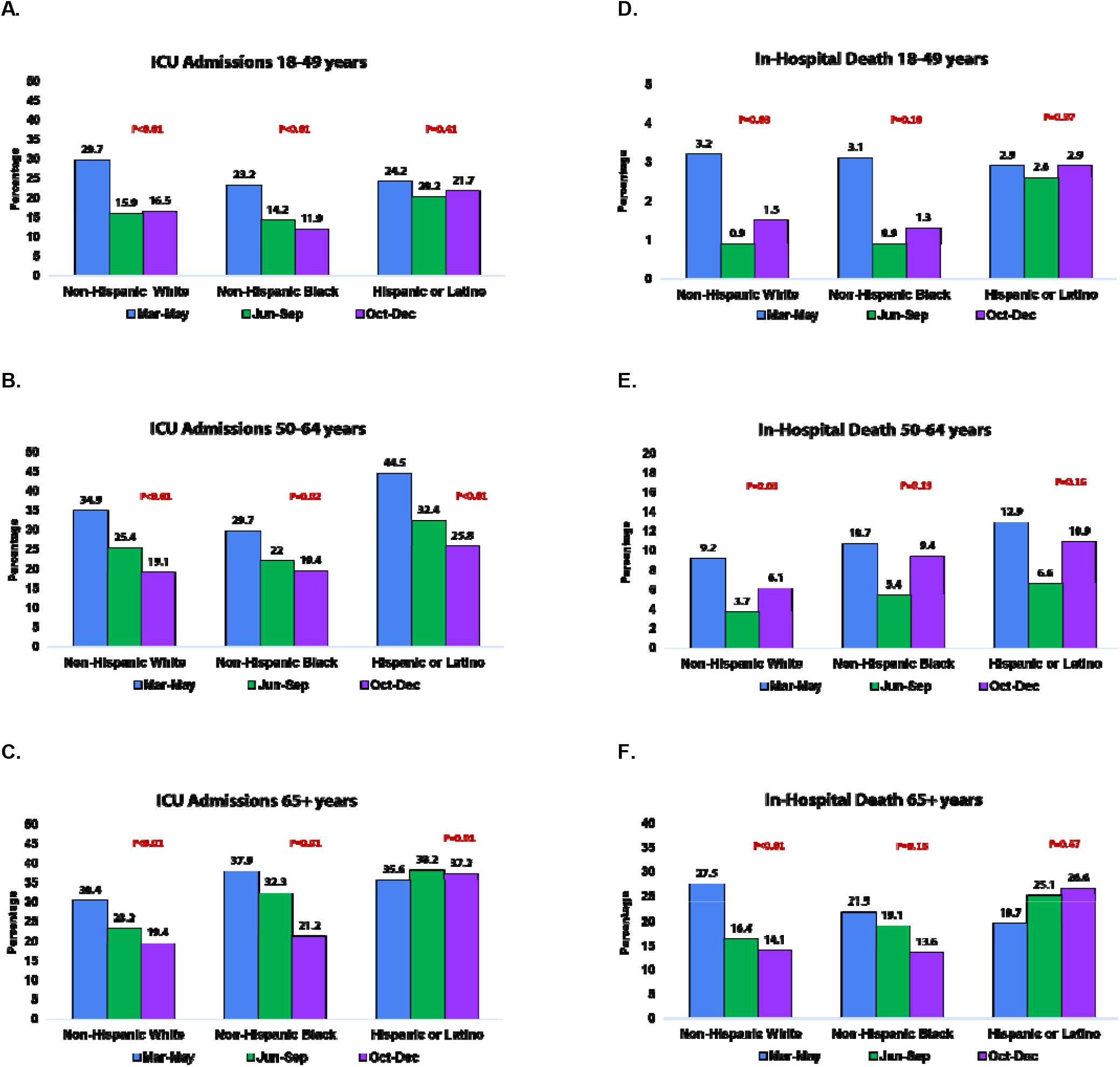
Trends^1^ in Percentage of Hospitalized Adults with COVID-19 with Severe Outcomes by Age and Race/ Ethnicity, COVID-NET, March-December 2020. 1 Logistic regression models were used to test the statistical significance of trends in outcomes over time by assigning monthly groupings (March-May, June-September, October-December) as a single continuous predictor in models for each age by race strata.

### Monthly Rates of Hospitalizations, ICU admissions and In-Hospital Deaths

There were 3 distinct peaks in monthly rates of COVID-19-associated hospitalizations, ICU admissions and in-hospital deaths (Figure 4, Supplemental Table 1) in April, July and December 2020. The highest rates for all 3 outcomes occurred during December 2020 (hospitalizations 105.3 per 100,000 population; ICU admissions 20.2; and in-hospital deaths 11.7). While rates for all 3 outcomes increased with age, trends over time were similar across all 3 age groups (Supplemental Figure 1, Supplemental Table 1).

**Figure 4.**
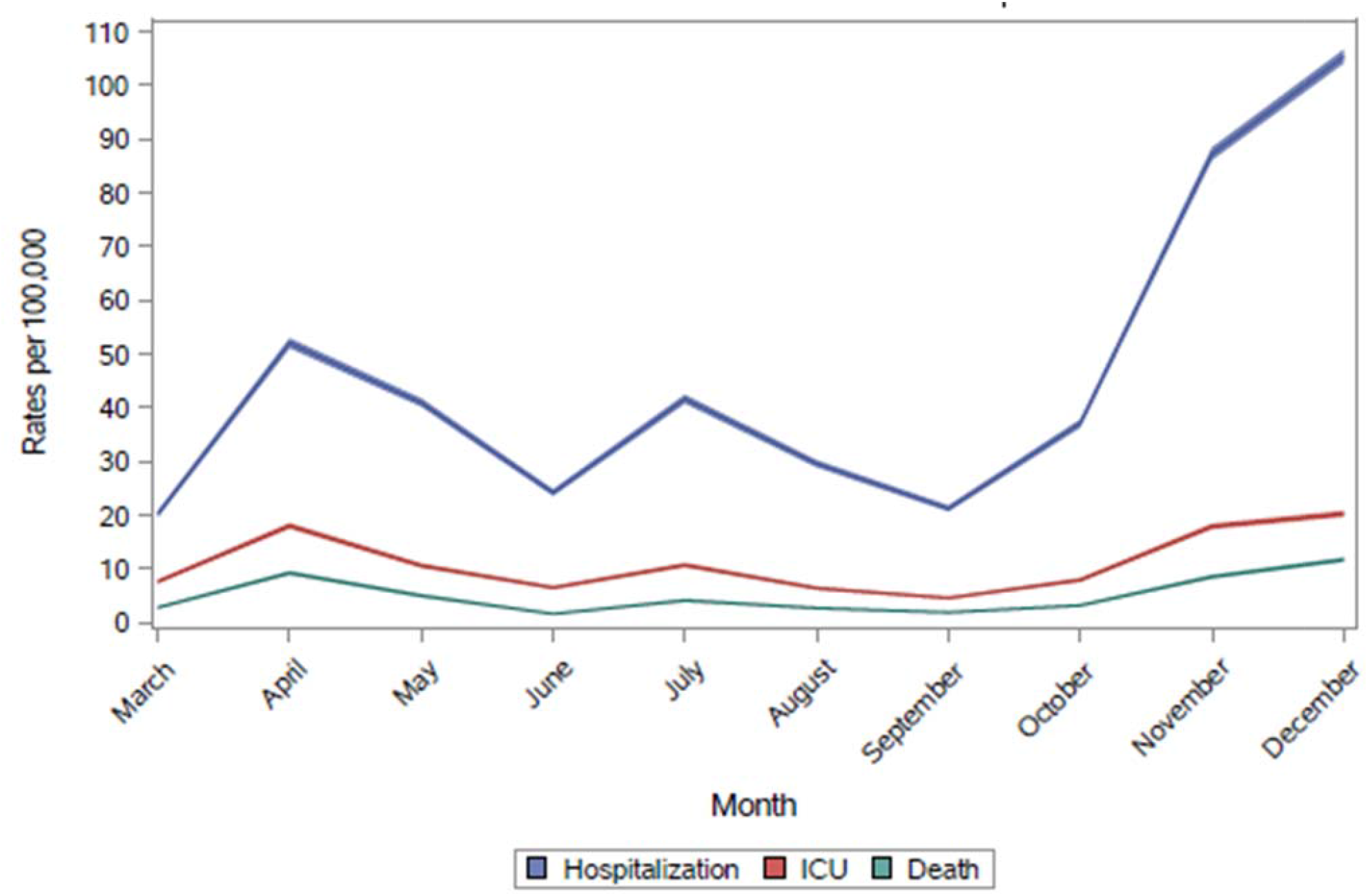
Rates (with 95% Confidence Intervals) of COVID-19- Associated Hospitalization^1^, Intensive Care Unit Admission^2^ and in-Hospital Death^1^ by Month, COVID-NET, March-December 2020. 1. Unadjusted hospitalization rates per 100,000 population were calculated by taking the total number of cases each month, divided by the National Center for Health Statistics’ vintage 2019 bridge-race postcensal population estimates for the counties included in surveillance. 2. Unadjusted ICU admission and death rates among hospitalized patients were calculated using the weighted number of sampled cases per month with each outcome as the numerator, divided by the National Center for Health Statistics’ vintage 2019 bridge-race postcensal population estimates for the counties included in surveillance.

## Discussion

Using a large, geographically diverse, population-based surveillance network for COVID-19-associated hospitalizations, we found that monthly rates of hospitalizations, ICU admissions and in-hospital deaths were highest in December, corresponding with the third peak of the U.S. pandemic. Among hospitalized cases, the percentage of ICU admissions, invasive mechanical ventilation, vasopressor support and in-hospital deaths declined during the first 7 months of the pandemic but remained stable or increased in November and December. While percentage of ICU admissions decreased among White and Black patients in all age groups over time, there were no statistically significant decreases in ICU admission percentages among Hispanic persons aged 18-49 or ≥65 years. In addition, we observed statistically significant decreases in the percentage of in-hospital deaths for White patients over time, but not for Black or Hispanic patients. Reasons for the observed trends in rates and percentages of interventions and outcomes are likely multifactorial and might be driven by patterns of community-level transmission, socioeconomic factors, and patient-, provider-and healthcare system-level factors.

Changes in community-level COVID-19 incidence was likely the largest contributor to observed trends in COVID-NET rates of hospitalizations, ICU admissions and in-hospital deaths (15); monthly changes in overall COVID-19 incidence corresponded with monthly changes in COVID-NET hospitalization rates (<u>CDC COVID Data Tracker</u>). Age likely also played a role; the median age of hospitalized adults steadily increased from a low of 56 years in June to 65 years in December. Similar age-related patterns were observed among COVID-19 cases nationally, where the median age declined from 46 years in May to 37 years in July and COVID-19 incidence was highest among persons aged 20-29 years during June-August 2020 (18). Multi-pronged mitigation measures, including mask use, social distancing and case investigations, may have contributed to decreased hospitalization rates during periods between peaks including June and September-October (19, 26). However, colder weather resulting in increased time spent indoors and holiday gatherings likely contributed to increased community transmission and a surge in hospitalizations in November and December (27, 28). With the availability of multiple COVID-19 vaccines in the U.S. starting in December 2020, and recommendations from the Advisory Committee on Immunization Practices to prioritize vaccination of certain high risk groups (29) and older adults (30), it is anticipated that COVID-19-associated hospitalization rates will decrease over time. This analysis provides an examination of trends in hospitalizations, interventions and outcomes prior to the large-scale availability of COVID-19 vaccines in the United States.

Despite the increase in rates of hospitalizations, ICU admissions and in-hospital deaths during the winter peak of the pandemic, we observed substantial decreases in the percentage of hospitalized cases who received aggressive interventions or died in-hospital during the first 7 months of COVID-NET surveillance, similar to other studies (11, 12, 15, 16). Evolving healthcare provider practices, guided by increasing knowledge and experience in managing patients with COVID-19, likely contributed to improved outcomes (11, 12, 16). While the percentage of patients receiving invasive mechanical ventilation decreased over time across all age groups, use of non-invasive respiratory support modalities, such as BIPAP, CPAP and HFNC increased over the same time-period. Several studies have shown improved outcomes with prone positioning (31) and use of non-invasive ventilatory support in lieu of invasive mechanical ventilation (11, 16, 32) for patients with COVID-19-associated acute respiratory failure. Decreasing use of invasive mechanical ventilation may have also lessened the need for ICU level care, as other respiratory support modalities could be delivered in non-ICU settings. Increasing use of more effective COVID-19-associated treatments may also have positively impacted outcomes. Dexamethasone has been shown to reduce 28-day mortality among patients requiring supplemental oxygen or invasive mechanical ventilation (22); use of systemic corticosteroids in our study increased from less than 20% in March to almost 75% in December 2020. Remdesivir, which has been shown to reduce time to clinical recovery in patients with severe COVID-19 (21), was received by less than 2% of patients in March but increased to over 50% by December 2020. Meanwhile the use of medications found to be ineffective for treatment of COVID-19 (33), such as hydroxychloroquine decreased substantially by June 2020.

Our analysis contributes to an expanding body of literature on racial and ethnic disparities related to COVID-19 (5, 8, 34-39). While the percentage of patients with ICU admissions and in-hospital deaths decreased over time for White patients across all age groups, similar trends were not consistently observed for Black and Hispanic patients. The reasons for these findings need to be explored further. Possible contributing factors, which we did not account for in this analysis, include differences in underlying comorbidities among different racial/ethnic groups, geographic differences in care practices which may correlate with the geographic distribution of different racial/ethnic groups, and long-standing inequities in social determinants of health with downstream effects on overall health of racial/ethnic minority groups (40). Other studies have similarly found increased COVID-19-associated morbidity and mortality among Black (39) and Hispanic (41, 42) persons. Ensuring equitable access to COVID-19 vaccine among groups at highest risk for COVID-19-associated complications, many of whom comprise racial and ethnic minority groups, will be essential to reducing COVID-19-associated morbidity and mortality in the United States.

Observed trends in interventions and outcomes may in part reflect a complex interplay between COVID-19 incidence and healthcare system capacity. As hospitalization rates peaked in different phases and geographic locations, ICU admission rates likely also increased. However, during peaks, the percentage of hospitalized patients receiving ICU admission or mechanical ventilation could have decreased or been capped due to scarcity of resources. Similarly, changes in timing of discharge and discharge disposition may in part have been influenced by bed capacity or the ability to discharge patients to sub-acute or chronic care facilities. In our study, almost 7% of patients 65 years and older were discharged to hospice. Among all patients, the percentage discharged to hospice increased from 1% in March to 4% in December 2020. Patterns in mortality may also be influenced by healthcare system capacity. One study conducted among 88 Department of Veterans Affairs hospitals found that strains on ICU capacity were associated with higher rates of COVID-19-associated ICU mortality (43). Linkage of COVID-NET data to facility-level data could allow for more direct examination of the impact of resource availability on COVID-19-associated interventions and outcomes. In addition, surveillance for post-discharge outcomes can provide a more complete picture of the out-of-hospital mortality burden among patients hospitalized with COVID-19.

This analysis has several limitations. Because COVID-NET covers approximately 10% of the U.S. population, our findings may not be generalizable to the entire country. This analysis presented data for the entire network and did not account for differences across sites; several factors, including peaks in hospitalization rates and the racial/ethnic distribution of cases varied by site. Since SARS-CoV-2 testing was conducted at the discretion of healthcare providers, COVID-NET may not have captured all COVID-19-associated hospitalizations. Changes in testing practices over the course of the pandemic may have influenced trends as sicker patients were more likely to be tested early in the pandemic when testing capacity was limited. The relative standard errors for some estimates were >0.3, particularly for interventions or outcomes with low prevalence, such as RRT or in-hospital deaths among patients aged 18-49 years. Lastly, sample sizes were not sufficient to fully explore differences in interventions and outcomes by race/ethnicity or to produce estimates for all racial/ethnic groups including AI/AN and Asian/PI patients. In future analyses, COVID-NET will combine additional months of data to characterize trends in severe outcomes among these racial/ethnic groups.

This analysis describes changes in characteristics, interventions and outcomes among U.S. adults hospitalized with COVID-19 over the first 10 months of the pandemic. Large declines in the percentage of patients with COVID-19-associated interventions and outcomes during the first seven months of the pandemic were encouraging, and likely reflected successful implementation of mitigation strategies as well as increasing healthcare provider knowledge, experience and tools to combat COVID-19. However, concerning increases in rates of hospitalization, ICU admission and in-hospital death emerged during the third pandemic peak. This study highlights ongoing racial and ethnic disparities in COVID-19-associated outcomes, as improvements in markers of disease severity over time were differentially observed across racial/ethnic groups. It will be important to follow trends in patient characteristics and outcomes over time in order to monitor impacts of clinical and public health interventions, including vaccination, and to ensure that progress is being made in closing the gap in racial and ethnic disparities related to COVID-19-associated outcomes in the United States.

## Data Availability

Publicly available data referred to in this analysis can be found at: https://gis.cdc.gov/grasp/covidnet/covid19_3.html

## Acknowledgements

Susan Brooks, MPH, Jeremy Roland, MPH, Roxanne Archer, MPH, Sherry Quach, BS, California Emerging Infections Program; James Meek, MPH, Paula Clogher, MPH, Danyel Olson, MPH, Hazal Kayalioglu, BS, Adam Misiorski, MPH, Christina Parisi, MPH, Maria Correa, MPH, Tessa Carter, MPH, Gaggan Brar, MD, Carol Lyons, MPH, Connecticut Emerging Infections Program, Yale School of Public Health; Emily Fawcett, MPH, Siyeh Gretzinger, MPH, Katelyn Ward, MPH, Jeremiah Williams, MPH, Jana Manning, MPH, Asmith Joseph, MPH, Allison Roebling, DVM, MPH, Stephanie Lehman, RN, BSN, Taylor Eisenstein, MPH, Gracie Chambers, Chris Bower, MPH, Andrew Revis, MPH, Dana Goodenough, MPH, Robin Dhonau, MPH, Sam Sefton, MPH, Georgia Emerging Infections Program, Georgia Department of Health, Atlanta Veterans Affairs Medical Center, Foundation for Atlanta Veterans Education and Research; Stepy Thomas, MSPH, Suzanne Segler, MPH, Grayson Kallas, BS, Amy Tunali, MPH, Georgia Emerging Infections Program, Georgia Department of Health, Division of Infectious Diseases, School of Medicine, Emory University; Kenzie Teno, MPH, Iowa Department of Public Health; Alicia Brooks, MPH, Maryland Department of Health; Jim Collins, MPH, Sam Hawkins, BS, Justin Henderson, MPH, Shannon Johnson, MPH, Val Tellez Nunez, BS, Alex Kohrman, MPH, Michigan Department of Health and Human Services; Austin Bell, MS, Kayla Bilski, BS, Emma Contestabile, BS, Kristen Ehresmann, MPH, Claire Henrichsen, BS, Emily Holodick, MPH, Lisa Nguyen, BS, Katherine Schleiss, MPH, Samantha Siebman, MPH, Richard Danila, MPH, PhD, Ruth Lynfield, MD, Minnesota Department of Health; Kathy M. Angeles, MPH, Emily B. Hancock, MS, Yadira Salazar-Sanchez, MPH, Meaghan Novi, MPH, Sarah A. Khanlian, MPH, Caroline Habrun, DVM, MPHTM, Melissa Christian, BUS, BA, Dominic Rudin, BS, Mayvilynne Poblete, MA, MPH, New Mexico Emerging Infections Program; Nancy Spina, MPH, Suzanne McGuire, MPH, Adam Rowe, BA, Kerianne Engesser, MPH, New York State Department of Health; Sophrena Bushey, MHS, Virginia Cafferky, BS, Christina Felsen, MPH, Maria Gaitan, RaeAnne Kurtz, BS, Christine Long, MPH, Kevin Popham, MPH, Marissa Tracy, MPH, University of Rochester School of Medicine and Dentistry; Nicole West, MPH, Ama Owusu-Dommey, MPH, Public Health Division, Oregon Health Authority; Kylie Seeley, MD, MPH, Oregon Health and Science University School of Medicine; Kathy Billings, MPH, Katie Dyer, Anise Elie, MPH, RN, BSN, Karen Leib, RN, Terri McMinn, Danielle Ndi, MPH, Manideepthi Pemmaraju, BBS, MPH, John Ujwok, MPH, Tiffanie Markus, PhD, Vanderbilt University Medical Center; Melanie Crossland, MPH, Andrea George, MPH, Andrea Price, LPN, Ashley Swain, CHES, Laine McCullough, MPH, Jake Ortega, MPH, Ilene Risk, MPA, Ian Buchta, MPH, Tyler Riedesel, MPH, Andrew Haraghey, Caitlin Shaw, Amanda Carter, Salt Lake County Health Department; Keegan McCaffrey, Utah Department of Health; Mimi Huynh, MPH, Council of State and Territorial Epidemiologists; Rainy Henry, Sonja Mali Nti-Berko, MPH, Robert W. Pinner, MD, Centers for Disease Control and Prevention

---

**Supplemental Table 1.** Rates (and 95% Confidence Internals) of Hospitalization^1^, Intensive Care Unit Admission^2^ and In-Hospital Death^2^ per 100,000 Persons by Age Group and Month, COVID-NET, March-December 2020.

| Month | Overall |  |  | 18-49 Years |  |  | 50-64 Years |  |  | ≥65 Years |  |  |
| --- | --- | --- | --- | --- | --- | --- | --- | --- | --- | --- | --- | --- |
|  | Hospital-ization Rate (95% CI) | ICU Rate (95% CI) | In-Hospital Death Rate (95% CI) | Hospital-ization Rate (95% CI) | ICU Rate (95% CI) | In-Hospital Death Rate (95% CI) | Hospital-ization Rate (95% CI) | ICU Rate (95% CI) | In-Hospital Death Rate (95% CI) | Hospital-ization Rate (95% CI) | ICU Rate (95% CI) | In-Hospital Death Rate (95% CI) |
| **March** | 20.2 (19.6-20.8) | 7.6 (7.3-8.0) | 2.8 (2.6-3.0) | 8.7 (8.2-9.2) | 2.4 (2.1-2.6) | 0.2 (0.2-0.3) | 27.3 (26.0-28.6) | 11.1 (10.3-11.9) | 2.8 (2.4-3.2) | 45.4 (43.5-47.3) | 18.9 (17.7-20.1) | 10.6 (9.7-11.5) |
| **April** | 52.0 (51.1-52.9) | 18.0 (17.5-18.5) | 9.2 (8.9-9.6) | 21.2 (20.4-22.0) | 6.4 (6.0-6.8) | 0.9 (0.7-1.0) | 61.9 (59.9-63.9) | 22.4 (21.2-23.6) | 6.7 (6.1-7.4) | 131.4 (128.1-134.6) | 47.2 (45.3-49.2) | 37.4 (35.7-39.1) |
| **May** | 40.9 (40.2-41.7) | 10.6 (10.2-11.0) | 5.0 (4.7-5.3) | 21.5 (20.7-22.2) | 4.0 (3.7-4.3) | 0.4 (0.3-0.5) | 47.8 (46.0-49.5) | 14.6 (13.7-15.6) | 5.0 (4.5-5.6) | 90.4 (87.7-93.1) | 24.9 (23.5-26.3) | 18.8 (17.6-20.0) |
| **June** | 24.2 (23.6-24.8) | 6.5 (6.2-6.8) | 1.6 (1.5-1.8) | 15.7 (15.1-16.4) | 2.8 (2.5-3.1) | 0.2 (0.2-0.3) | 28.1 (26.8-29.4) | 8.3 (7.6-9.0) | 1.8 (1.5-2.2) | 44.3 (42.4-46.2) | 15.2 (14.1-16.3) | 5.5 (4.8-6.2) |
| **July** | 41.6 (40.8-42.4) | 10.7 (10.3-11.1) | 4.1 (3.9-4.4) | 25.6 (24.8-26.4) | 4.4 (4.1-4.8) | 0.5 (0.4-0.6) | 49.1 (47.4-50.8) | 14.3 (13.4-15.3) | 3.4 (3.0-3.9) | 79.6 (77.1-82.1) | 24.6 (23.2-26.0) | 16.0 (14.9-17.2) |
| **August** | 29.6 (28.9-30.2) | 6.4 (6.1-6.7) | 2.7 (2.5-2.9) | 17.7 (17.0-18.4) | 3.0 (2.7-3.3) | 0.2 (0.1-0.2) | 34.1 (32.7-35.6) | 8.1 (7.4-8.8) | 1.5 (1.2-1.8) | 59.2 (57.0-61.4) | 14.4 (13.3-15.4) | 11.7 (10.8-12.7) |
| **September** | 21.2 (20.7-21.8) | 4.6 (4.3-4.8) | 1.9 (1.7-2.0) | 11.5 (11.0-12.1) | 1.8 (1.6-2.0) | 0.3 (0.2-0.3) | 24.3 (23.0-25.5) | 5.7 (5.1-6.3) | 0.8 (0.6-1.0) | 46.4 (44.5-48.4) | 11.4 (10.4-12.3) | 8.0 (7.2-8.8) |
| **October** | 37.1 (36.3-37.8) | 7.9 (7.6-8.3) | 3.2 (3.0-3.4) | 17.4 (16.7-18.1) | 2.4 (2.1-2.6) | 0.3 (0.2-0.4) | 42.7 (41.0-44.3) | 9.6 (8.9-10.4) | 2.9 (2.5-3.3) | 88.8 (86.1-91.4) | 22.4 (21.1-23.8) | 12.2 (11.2-13.2) |
| **November** | 87.2 (86.1-88.4) | 17.9 (17.4-18.4) | 8.5 (8.2-8.9) | 34.7 (33.7-35.6) | 5.7 (5.3-6.1) | 0.7 (0.6-0.9) | 95.3 (92.9-97.7) | 20.9 (19.7-22.0) | 6.3 (5.7-7.0) | 234.0 (229.7-238.4) | 50.5 (48.4-52.5) | 34.8 (33.1-36.5) |
| **December** | 105.3 (104.1-106.6) | 20.2 (19.7-20.8) | 11.7 (11.3-12.1) | 39.5 (38.5-40.5) | 7.0 (6.6-7.5) | 0.9 (0.7-1.0) | 111.9 (109.2-114.5) | 20.8 (19.6-21.9) | 9.7 (9.0-10.5) | 293.8 (289.0-298.7) | 58.9 (56.8-61.1) | 46.6 (44.7-48.6) |

**Supplemental Table 2:** Demographic Characteristics of Sampled versus All Cases Hospitalized with COVID-19, COVID-NET, March-December 2020.

| Variable | Sampled Cases; Unweighted N (Wtd %) | Total Cases; Unweighted N (UnWtd %) | Mar Sampled Wtd % | Mar Total UnWtd % | Apr Sampled Wtd % | Apr Total UnWtd % | May Sampled Wtd % | May Total UnWtd % | Jun Sampled Wtd % | June Total UnWtd % | Jul Sampled Wtd % | Jul Total UnWtd % |
| --- | --- | --- | --- | --- | --- | --- | --- | --- | --- | --- | --- | --- |
| **Median Age** | 18,508 (62.1) | 116,743 (62) | 61 | 61 | 63 | 63 | 60 | 60 | 55 | 56 | 58 | 57 |
| **Sex** |  |  |  |  |  |  |  |  |  |  |  |  |
| Male | 9,410 (51.1) | 59,131 (50.7) | 59 | 55 | 53 | 53 | 49 | 50 | 51 | 49 | 47 | 50 |
| Female | 90,98 (48.9) | 57,608 (49.3) | 42 | 45 | 47 | 47 | 51 | 50 | 49 | 51 | 53 | 51 |
| **Race/Ethnicity** |  |  |  |  |  |  |  |  |  |  |  |  |
| NH White | 7,469 (41.7) | 46,923 (40.8) | 35 | 37 | 39 | 36 | 31 | 32 | 26 | 27 | 29 | 29 |
| NH Black | 4,374 (27.2) | 31,019 (26.9) | 43 | 40 | 32 | 34 | 30 | 29 | 30 | 30 | 38 | 37 |
| NH AI/AN | 398 (1.6) | 1619 (1.4) | 0 | 1 | 1 | 1 | 2 | 2 | 2 | 2 | 1 | 1 |
| NH Asian | 1,449 (5.3) | 5793 (5) | 5 | 5 | 4 | 5 | 5 | 5 | 7 | 6 | 5 | 5 |
| NH Multiracial | 67 (0.4) | 377 (0.3) | 0 | 0 | 0 | 0 | 0 | 0 | 1 | 0 | 1 | 1 |
| Hispanic or Latino | 4,192 (20.5) | 23,079 (20) | 13 | 14 | 21 | 21 | 29 | 27 | 32 | 30 | 24 | 24 |
| Unknown | 556 (3.2) | 6313 (5.5) | 3 | 4 | 2 | 3 | 2 | 4 | 3 | 5 | 3 | 4 |
| **Surveillance Site** |  |  |  |  |  |  |  |  |  |  |  |  |
| CA | 937 (7.6) | 8923 (7.6) | 6 | 6 | 4 | 4 | 3 | 4 | 12 | 9 | 11 | 11 |
| CO | 1,024 (9.7) | 11,206 (9.6) | 17 | 17 | 14 | 13 | 6 | 8 | 6 | 5 | 4 | 5 |
| CT | 1,021 (4.9) | 5708 (4.9) | 7 | 8 | 12 | 12 | 7 | 7 | 2 | 2 | 1 | 1 |
| GA | 955 (15.2) | 17,478 (15) | 24 | 24 | 9 | 10 | 9 | 9 | 18 | 22 | 35 | 33 |
| IA | 159 (0.5) | 701 (0.6) | 0 | 0 | 1 | 1 | 1 | 1 | 0 | 0 | 1 | 0 |
| MD | 1,436 (22.1) | 25,909 (22.2) | 18 | 18 | 34 | 34 | 38 | 38 | 24 | 25 | 15 | 16 |
| MI | 552 (3.4) | 3880 (3.3) | 7 | 7 | 5 | 5 | 1 | 1 | 0 | 1 | 1 | 1 |
| MN | 5,849 (10.6) | 12,476 (10.7) | 3 | 3 | 6 | 6 | 15 | 14 | 11 | 10 | 7 | 7 |
| NM | 641 (4.3) | 5107 (4.4) | 1 | 1 | 2 | 2 | 3 | 3 | 3 | 3 | 4 | 4 |
| NYA | 303 (2.2) | 2522 (2.2) | 3 | 3 | 3 | 2 | 2 | 2 | 1 | 1 | 1 | 1 |
| NYR | 446 (3.5) | 4052 (3.5) | 3 | 3 | 2 | 2 | 4 | 4 | 2 | 2 | 1 | 1 |
| OH | 748 (3.5) | 4043 (3.5) | 3 | 3 | 3 | 3 | 4 | 4 | 5 | 5 | 5 | 5 |
| OR | 719 (2.4) | 2861 (2.5) | 3 | 3 | 2 | 2 | 1 | 1 | 2 | 2 | 2 | 2 |
| TN | 1869 (7) | 8045 (6.9) | 6 | 6 | 3 | 3 | 4 | 4 | 10 | 8 | 9 | 9 |
| UT | 1,849 (3.3) | 3832 (3.3) | 1 | 1 | 1 | 1 | 2 | 2 | 5 | 5 | 4 | 4 |

\*Wtd = weighted percentage; Unwtd= unweighted percentage

Supplemental Table 2 Continued...
| Variable | Aug<br>Sampled<br>Wtd % | Aug Total<br>UnWtd % | Sept<br>Sampled<br>Wtd % | Sept Total<br>UnWtd % | Oct<br>Sampled<br>Wtd % | Oct Total<br>UnWtd % | Nov<br>Sampled<br>Wtd % | Nov Total<br>UnWtd % | Dec<br>Sampled<br>Wtd % | Dec Total<br>UnWtd % |
| --- | --- | --- | --- | --- | --- | --- | --- | --- | --- | --- |
| **Median Age** | 60 | 58 | 60 | 60 | 62 | 62 | 64 | 64 | 65 | 65 |
| **Sex** |  |  |  |  |  |  |  |  |  |  |
| Male | 49 | 49 | 49 | 49 | 53 | 51 | 50 | 51 | 53 | 50 |
| Female | 51 | 51 | 51 | 52 | 47 | 49 | 50 | 49 | 48 | 50 |
| **Race/Ethnicity** |  |  |  |  |  |  |  |  |  |  |
| NH White | 35 | 33 | 41 | 41 | 45 | 46 | 51 | 50 | 51 | 48 |
| NH Black | 33 | 32 | 26 | 25 | 22 | 21 | 19 | 19 | 23 | 23 |
| NH AI/AN | 1 | 1 | 1 | 1 | 1 | 1 | 3 | 2 | 2 | 2 |
| NH Asian | 7 | 7 | 8 | 7 | 7 | 5 | 6 | 5 | 4 | 5 |
| NH Multiracial | 0 | 0 | 0 | 1 | 1 | 0 | 0 | 0 | 1 | 0 |
| Hispanic or Latino | 21 | 22 | 21 | 22 | 21 | 22 | 17 | 18 | 17 | 15 |
| Unknown | 3 | 5 | 4 | 5 | 4 | 5 | 4 | 8 | 3 | 7 |
| **Surveillance Site** |  |  |  |  |  |  |  |  |  |  |
| CA | 13 | 14 | 8 | 12 | 5 | 5 | 4 | 5 | 11 | 11 |
| CO | 5 | 4 | 5 | 6 | 13 | 13 | 15 | 14 | 8 | 7 |
| CT | 2 | 1 | 2 | 2 | 3 | 3 | 5 | 5 | 5 | 5 |
| GA | 25 | 25 | 18 | 17 | 12 | 12 | 8 | 8 | 13 | 13 |
| IA | 1 | 1 | 1 | 1 | 1 | 1 | 1 | 1 | 0 | 0 |
| MD | 19 | 19 | 23 | 21 | 17 | 17 | 18 | 18 | 18 | 19 |
| MI | 1 | 1 | 2 | 2 | 4 | 4 | 5 | 5 | 4 | 4 |
| MN | 12 | 12 | 13 | 13 | 14 | 14 | 14 | 15 | 9 | 9 |
| NM | 2 | 3 | 3 | 3 | 7 | 7 | 7 | 7 | 5 | 5 |
| NYA | 1 | 1 | 1 | 1 | 2 | 2 | 2 | 2 | 4 | 4 |
| NYR | 1 | 1 | 2 | 2 | 2 | 2 | 4 | 4 | 7 | 7 |
| OH | 4 | 4 | 5 | 5 | 4 | 4 | 3 | 3 | 2 | 2 |
| OR | 3 | 3 | 3 | 4 | 3 | 3 | 3 | 3 | 3 | 2 |
| TN | 8 | 8 | 8 | 8 | 9 | 9 | 7 | 7 | 8 | 8 |
| UT | 3 | 3 | 6 | 6 | 6 | 6 | 4 | 4 | 3 | 3 |

\*Wtd = weighted percentage; Unwtd= unweighted percentage

**Supplemental Table 3.** Percentage Receiving COVID-19 Associated Treatments Among Hospitalized Adults with COVID-19 by Month, COVID-NET, March-December 2020.

| Month | Unweighted N | Cortico-steroids | Remdesivir | Azithromycin | Convalescent Plasma | HCQ<sup>1</sup> | Tocilizumab | Zinc | Vitamin C | Protease Inhibitor<sup>2</sup> | Other Drug<sup>3</sup> | Study Drug<sup>4</sup> | Remdesivir Study |
| --- | --- | --- | --- | --- | --- | --- | --- | --- | --- | --- | --- | --- | --- |
| Weighted % |  |  |  |  |  |  |  |  |  |  |  |  |  |
| Overall | 18,508 | 54.80 | 36.13 | 25.96 | 10.48 | 8.19 | 2.54 | 6.86 | 6.61 | 0.30 | 1.64 | 0.72 | 0.29 |
| March | 1,375 | 18.88 | 1.75 | 43.52 | 0.46 | 48.93 | 4.90 | 3.61 | 2.55 | 4.28 | 0.94 | 0.06 | 1.44 |
| April | 3,488 | 20.87 | 2.78 | 32.84 | 3.83 | 40.85 | 7.83 | 7.99 | 7.11 | 0.60 | 0.97 | 0.37 | 0.40 |
| May | 3,466 | 19.59 | 15.53 | 22.67 | 10.40 | 8.58 | 6.56 | 8.94 | 8.91 | 0.09 | 0.94 | 1.28 | 0.30 |
| June | 1,523 | 31.17 | 29.88 | 22.02 | 12.42 | 0.56 | 3.67 | 6.20 | 6.46 | 0.02 | 0.52 | 2.50 | 0.36 |
| July | 2,176 | 61.12 | 31.61 | 29.95 | 13.97 | 0.78 | 2.48 | 6.12 | 5.94 | 0.07 | 1.01 | 1.41 | 0.02 |
| August | 1,823 | 59.19 | 32.06 | 26.02 | 11.64 | 1.09 | 0.87 | 5.81 | 5.88 | 0.34 | 0.85 | 1.01 | 0.34 |
| September | 1,472 | 58.57 | 38.74 | 22.58 | 11.28 | 0.24 | 1.07 | 5.91 | 5.28 | 0.00 | 1.18 | 1.03 | 0.80 |
| October | 901 | 65.00 | 45.47 | 24.14 | 17.42 | 0.38 | 1.38 | 7.75 | 6.56 | 0.00 | 1.08 | 0.55 | 0.13 |
| November | 1,429 | 73.98 | 54.33 | 24.07 | 12.18 | 0.39 | 0.60 | 5.23 | 5.31 | 0.00 | 1.17 | 0.58 | 0.26 |
| December | 855 | 74.19 | 53.82 | 22.19 | 9.71 | 0.36 | 0.36 | 8.15 | 8.22 | 0.00 | 3.85 | 0.19 | 0.05 |

HCQ= Hydroxychloroquine
1. Hydroxychloroquine with or without azithromycin
2. Protease inhibitors excluded if patient had known history of HIV
3. Other drugs include: chloroquine, sarilumab (Kevzara), Baricitinib (Olmiant), losartan, immunomodulators, casirivimab/imdevimab (REGN-COV2), bamlanivimab, IVIG, interferon, ivermectin, cholecalciferol and daclizumab
4. Study drugs other than remdesivir studies include: hydroxychloroquine study, losartan study, baricitinib study, monoclonal antibody study, interferon study, sarilumab (Kevzara) study, tocilizumab study, casirivimab/Imdevimab (REGN-COV2) study, kinase Inhibitor study, convalescent plasma study, dexamethasone study and other specified study drugs

**Supplemental Table 4.**
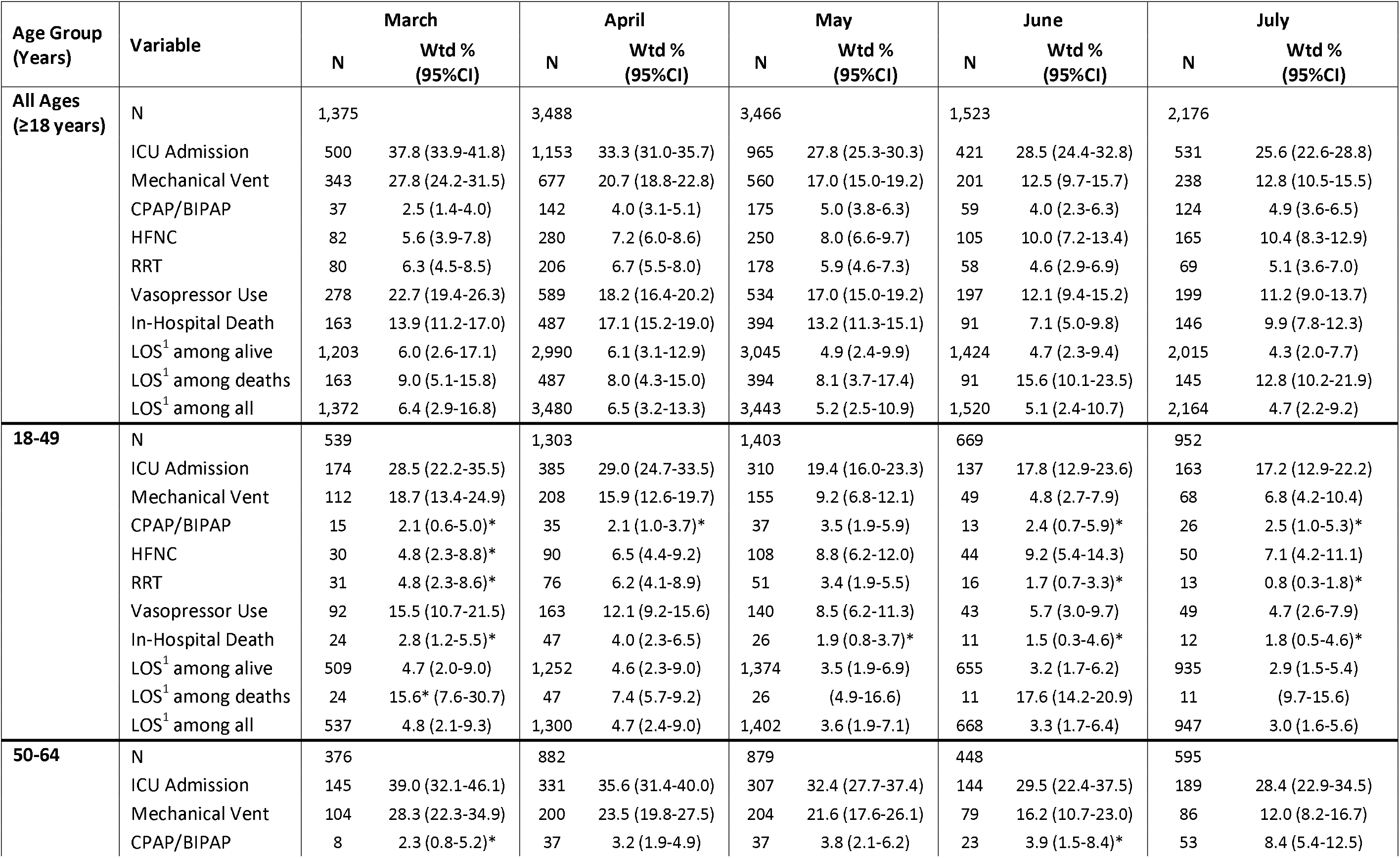
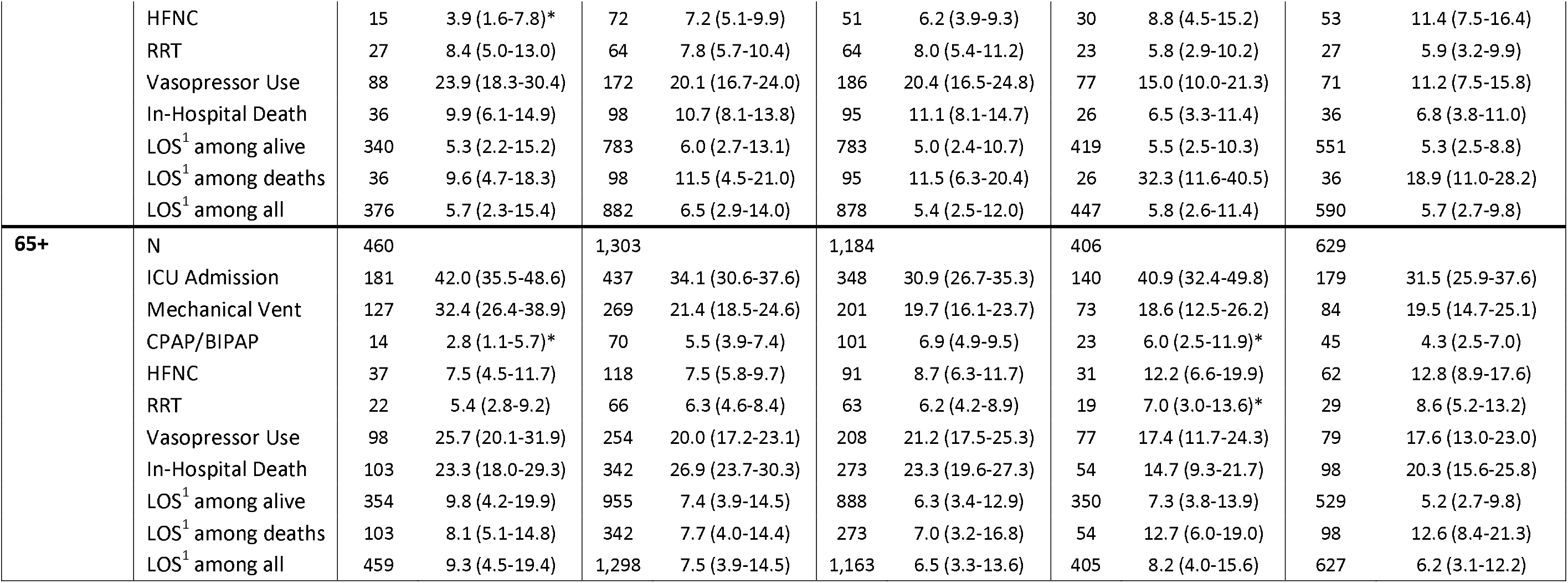
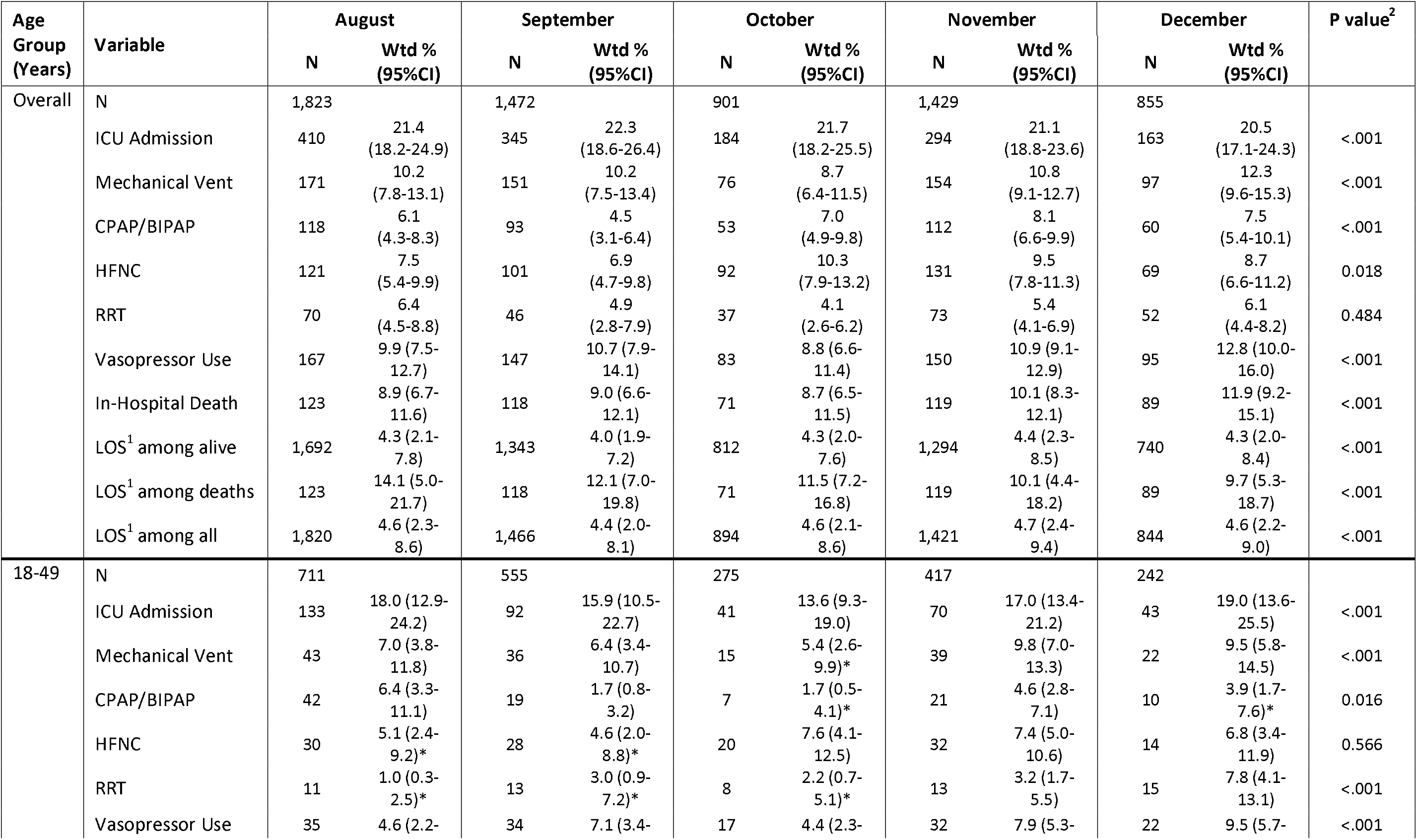
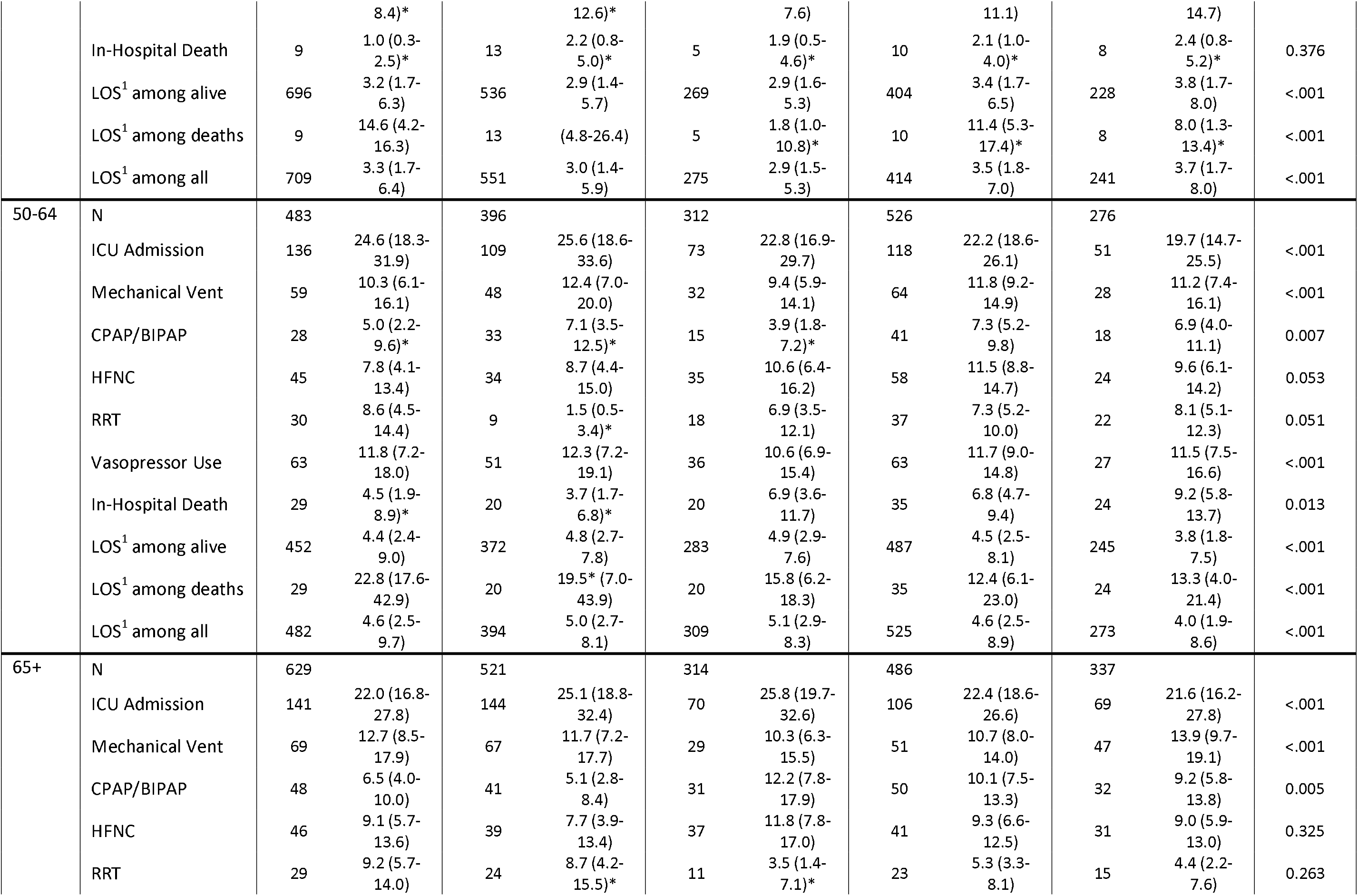
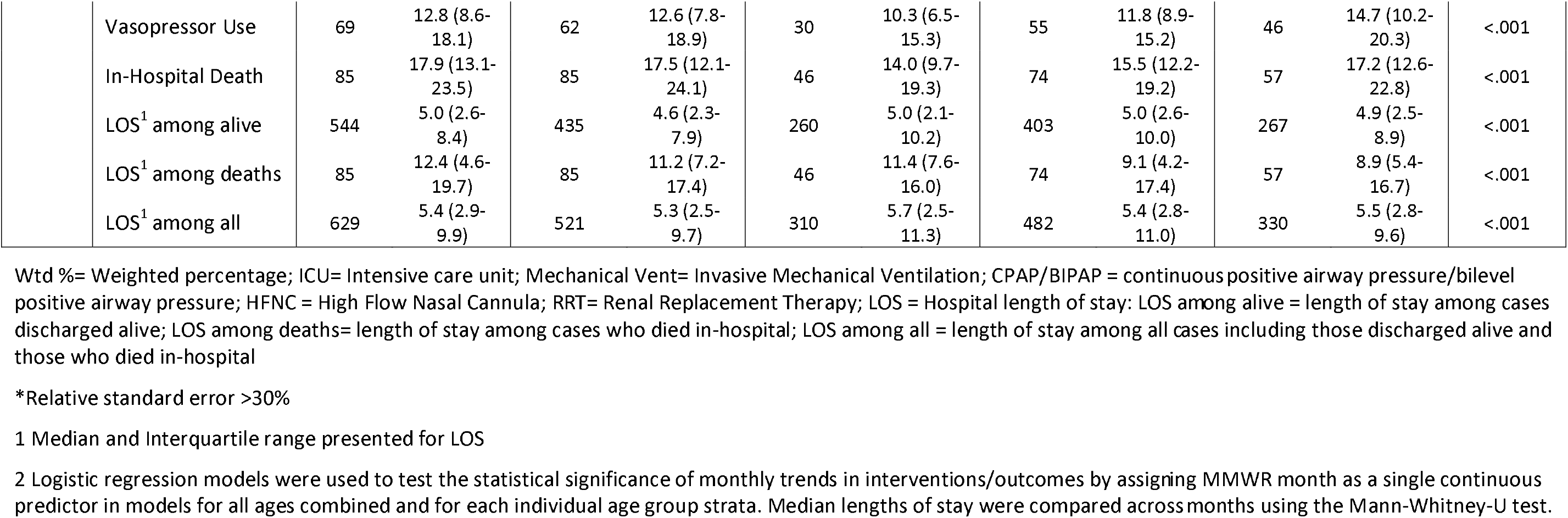
Percentage with Clinical Interventions and Outcomes among Adults with Laboratory-Confirmed COVID-19-Associated Hospitalizations by Month, COVID-NET, March-December 2020.

**Supplemental Table 5.**
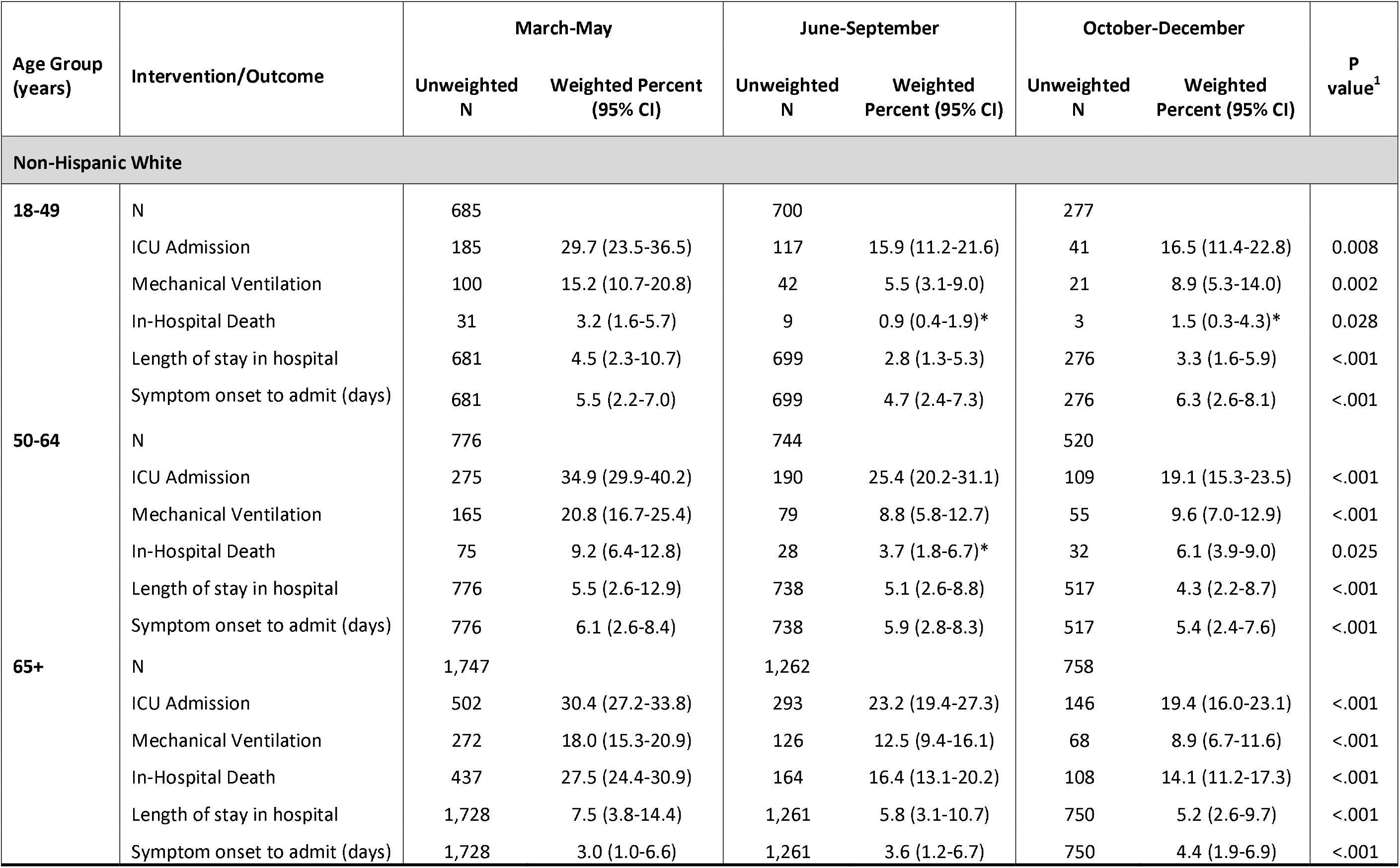
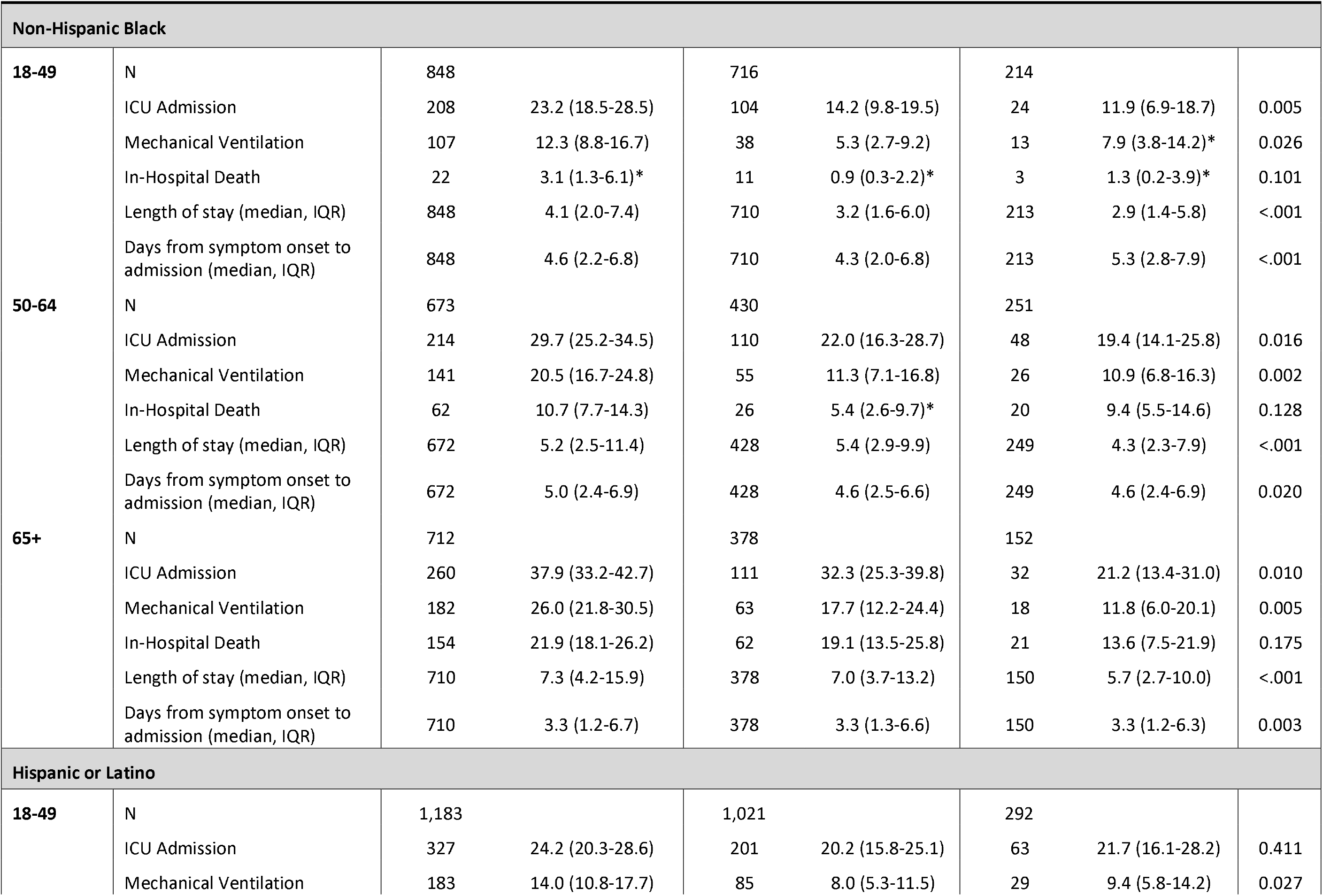
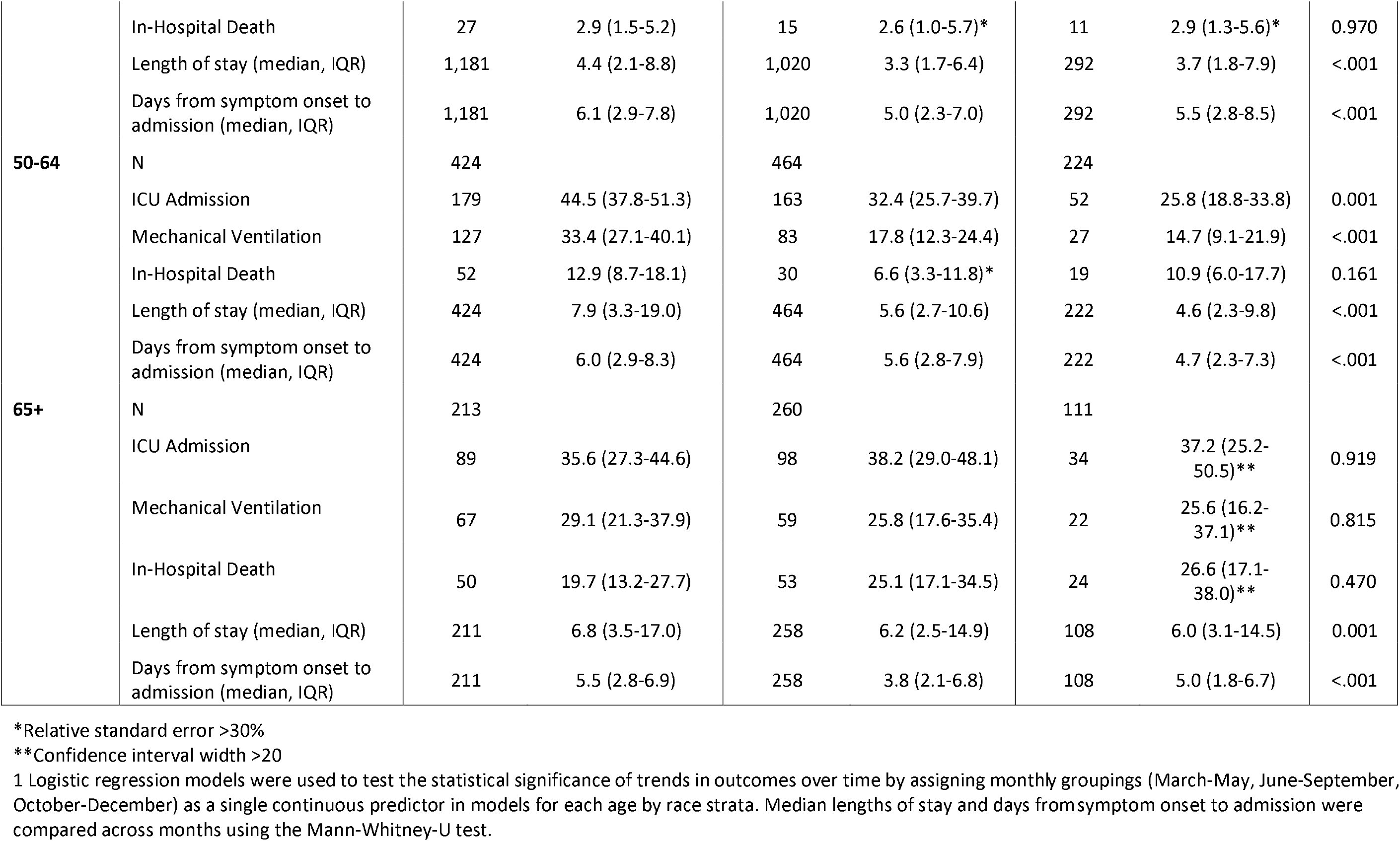
Percentage with Clinical Interventions and Outcomes among Adults with Laboratory-Confirmed COVID-19-Associated Hospitalizations by Race and Ethnicity, Age Group and Quarter, COVID-NET, March-December 2020.

**Supplemental Figure 1.**
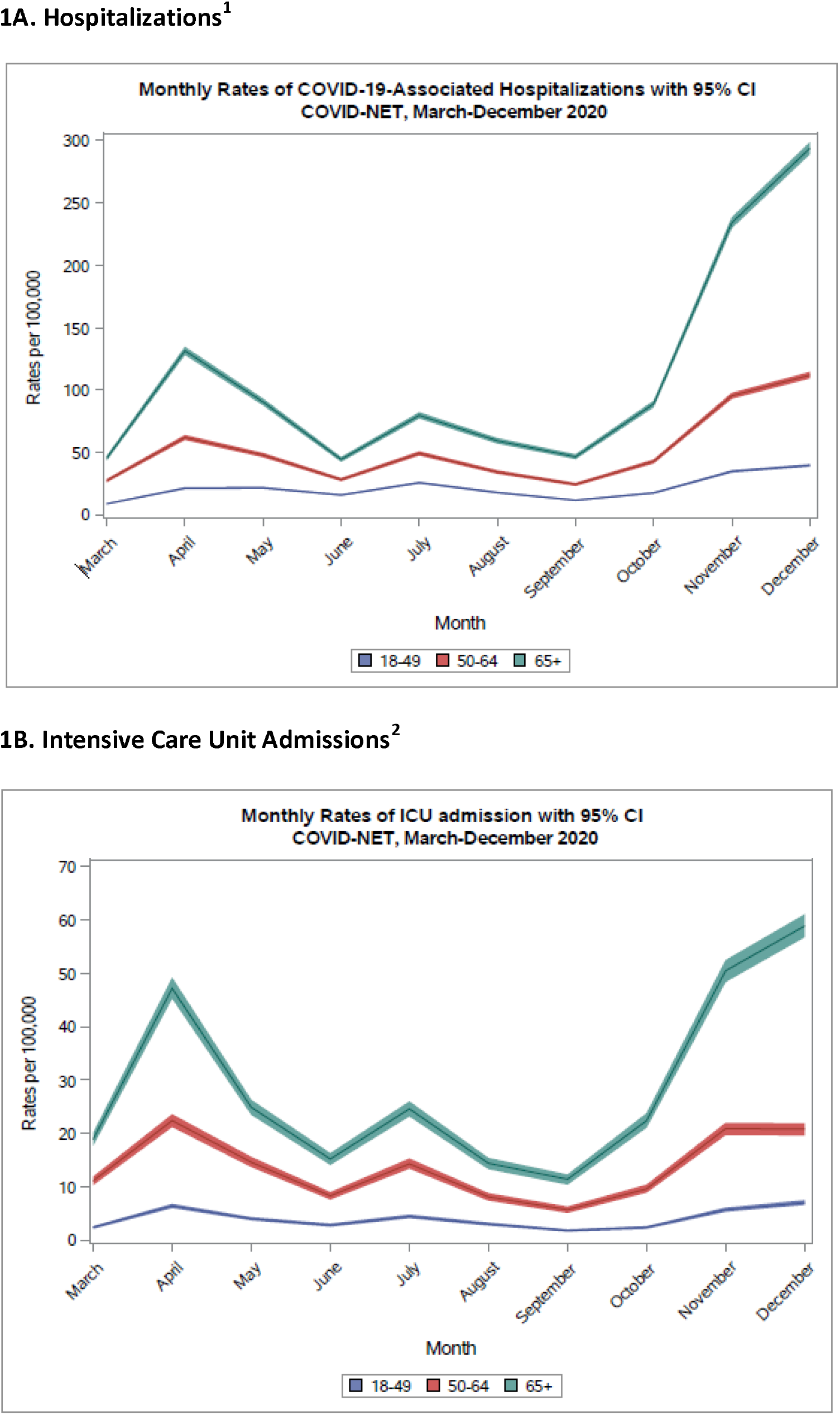
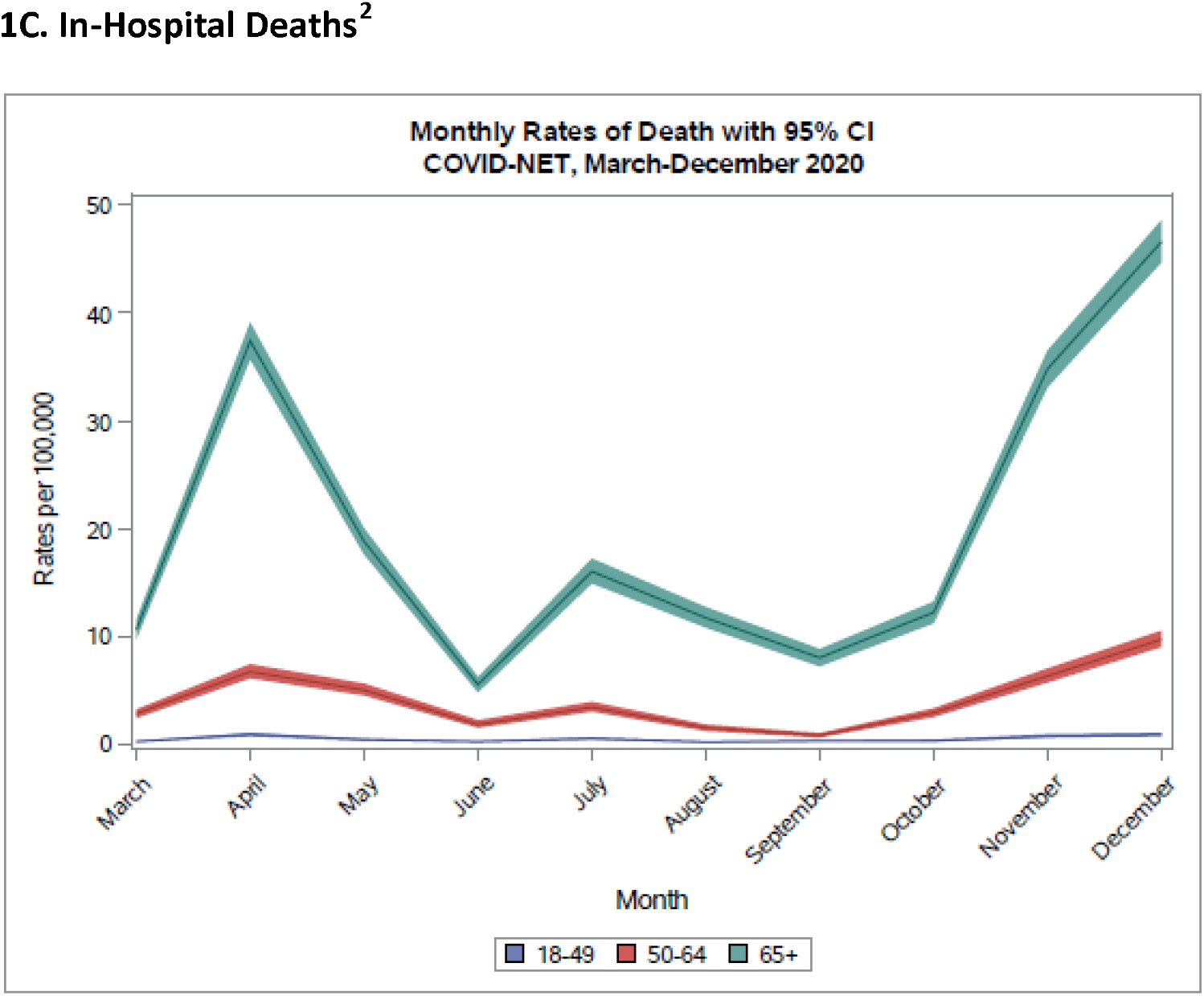
Rates of Hospitalization, Intensive Care Unit Admission and In-Hospital Death by Age Group and Month, COVID-NET, March-December 2020. 1. Unadjusted hospitalization rates per 100,000 population were calculated by taking the total number of cases each month, divided by the National Center for Health Statistics’ vintage 2019 bridge-race postcensal population estimates for the counties included in surveillance. 2. Unadjusted ICU admission and death rates among hospitalized patients were calculated using the weighted number of sampled cases per month with each outcome as the numerator, divided by the National Center for Health Statistics’ vintage 2019 bridge-race postcensal population estimates for the counties included in surveillance.

